# Low Vision Rehabilitation Training and Referral Patterns Among Ophthalmologists

**DOI:** 10.1101/2022.03.15.22272453

**Authors:** Joshua L. Robinson, Ansley Kunnath, Janice C. Law

## Abstract

**Objective:** Vision impairment represents a growing burden to society and training protocols related to low vision rehabilitation (vision rehab) vary across ophthalmology residency programs. We surveyed practicing ophthalmologists regarding their vision rehab knowledge, confidence levels, and referral thresholds. We categorized subjects and compared response patterns between groups.

**Design:** Prospective observational

**Subjects:** 185 practicing ophthalmologists

**Methods:** We created an Ophthalmology Low Vision Questionnaire and administered it to all enrolled subjects via email. We categorized subjects based on duration of practice, subspecialty area, and exposure to vision rehab during residency training. We drew conclusions by comparing responses between various subject categories.

**Main Outcome Measures:** The primary outcome measure was comparison of confidence levels and thresholds for vision rehab referral across groups. We used statistical tests to look for associations between practice duration, subspecialty area, vision rehab exposure during residency, and referral patterns.

**Results:** Ophthalmologists practicing for 6 or less years were more likely to have had a formal vision rehab rotation during their residency training compared to ophthalmologists practicing for 7 or more years (*P* = 0.03). Ophthalmologists who completed a formal vision rehab rotation during residency reported greater confidence in their ability to appropriately identify patients who could benefit from vision rehab referral (*P* = 0.04) and referred patients at earlier visual acuity thresholds (*P* = 0.04). Clinical subspecialty did not have a significant effect on vision rehab referral confidence or thresholds.

**Conclusions:** Vision rehab rotations during residency lead to improved referral confidence and earlier referral for these vital services. Standardization of vision rehab exposure across ophthalmology residency programs can help to improve outcomes for visually impaired patients.

## Introduction

Permanent visual impairment is a rapidly growing issue which represents a large burden to both the visually impaired individual and to society.^1–3^ In 2013, the total economic burden of eye disorders and vision loss in the United States was about $139 billion.^4^ A discipline known as low vision rehabilitation (vision rehab) aims to relieve that burden and allow visually impaired individuals to maximize their independence and quality of life.^5^

While provision of clinical vision rehab services is often undertaken by optometrists, ophthalmologists play an important role in identifying and referring patients who need these services. Within the past several years, the American Academy of Ophthalmology (Academy) has advocated for early referral to vision rehab programs as the standard of care for patients who suffer from irreversible vision loss.^6^ The Accreditation Council for Graduate Medical Education (ACGME) states in their Common Program Requirements that ophthalmology residents must have access to faculty members with expertise in vision rehab and must demonstrate competence in their knowledge of vision rehab.^7^ This broad guidance allows ophthalmology residency programs to remain highly variable in the amount of vision rehab exposure received by trainees.

This survey-based study aimed to assess practicing ophthalmologists’ levels of understanding, referral confidence, and utilization of vision rehab services. Analysis of these findings can increase awareness and understanding of vision rehab as a key treatment modality in caring for those with impaired vision, a population which is expected to continue to grow worldwide.^1^

## Methods

This study was approved by the Institutional Review Board at Vanderbilt University and followed the tenets of the Declaration of Helsinki. The study was approved with consent exemption.

We developed a survey (Appendix) to assess perceptions and practices related to vision rehab referrals among ophthalmologists. We invited practicing ophthalmologists, regardless of subspecialty area or duration of practice, to be included in the study. We excluded physicians currently in residency or fellowship training programs.

We enrolled subjects via Academy email groups and social media solicitation. We sent physicians who agreed to voluntary inclusion in the study a link to an anonymous RedCap survey via email. We drew conclusions through compilation and statistical analysis of the survey responses.

### Statistical Analysis

We analyzed data in groups based on subjects’ duration of practice, subspecialty practice area, and self-reported exposure during residency to vision rehab programs. We used chi-squared tests to compare the percentage of participants within a specific group that answered “yes” to each question. We used one-way ANOVA and two-tailed, unpaired t-tests to compare the mean confidence level and referral criteria of different groups.

## Results

A total of 185 ophthalmologists and one optometrist responded to this survey. The responses of the lone optometrist were excluded. Results for the 185 ophthalmologists surveyed are summarized in Table 1.

**Table 1.** Comparison of Low Vision Rehabilitation Training and Referrals by Duration of Practice.

| Years in Practice | < 3 years | 3 to 6 years | 7 to 10 years | > 10 years |
| --- | --- | --- | --- | --- |
| **Number of Subjects (%)** | 66 (36) | 70 (38) | 21 (11) | 28 (15) |
| **Residency Training** |  |  |  |  |
| Had VR rotation, No. (%) | 38 (58) | 36 (51) | 8 (38) | 10 (36) |
| Had OT or CLVTs, No. (%)<sup>a</sup> | 52 (85) | 44 (69) | 15 (75) | 13 (52) |
| **VR Referral** |  |  |  |  |
| Has referred to VR, No. (%) | 54 (82) | 65 (93) | 19 (90) | 26 (93) |
| Confidence from 1-5 (mean) | 3.71 | 3.66 | 3.67 | 3.89 |
| **Indication for VR Referral<sup>b</sup>** |  |  |  |  |
| Reduced BCVA, No. (%) | 62 (94) | 67 (96) | 21 (100) | 27 (96) |
| Visual field loss, No. (%) | 59 (89) | 64 (91) | 17 (81) | 24 (86) |
| Decreased CS, No. (%) | 32 (48) | 25 (36) | 7 (33) | 9 (32) |
| Other, No. (%) | 8 (12) | 8 (11) | 2 (10) | 2 (7) |
| **BCVA Threshold for VR Referral<sup>c</sup>** |  |  |  |  |
| 20/25 to 20/63, No. (%) | 11 (18) | 17 (25) | 5 (24) | 10 (37) |
| 20/80 to 20/160, No. (%) | 32 (52) | 33 (49) | 7 (33) | 12 (44) |
| 20/200 to 20/400, No. (%) | 18 (29) | 17 (25) | 8 (38) | 4 (15) |
| worse than 20/400, No. (%) | 1 (2) | 0 (0) | 1 (5) | 1 (4) |
| **VR Resources Presented<sup>a</sup>** |  |  |  |  |
| VR services within clinic, No. (%) | 25 (38) | 17 (24) | 8 (38) | 19 (68) |
| Community VR providers, No. (%) | 33 (50) | 50 (71) | 14 (67) | 13 (46) |
| State-based resources, No. (%) | 30 (45) | 29 (41) | 8 (38) | 8 (29) |
| Non-governmental org, No. (%) | 20 (30) | 20 (29) | 7 (33) | 8 (29) |
| Online or AAO resources, No. (%) | 8 (12) | 11 (16) | 0 (0) | 8 (29) |
| Other, No. (%) | 3 (5) | 1 (1) | 1 (5) | 0 (0) |

<sup>a</sup>subjects who responded "unsure" (n = 15) were excluded from percentage calculations
<sup>b</sup>some subjects responded "yes" to multiple options
<sup>c</sup>subjects who did not respond (n = 8) were excluded from percentage calculations
VR: vision rehabilitation
OT: occupational therapist
CLVT: certified low vision therapist
BCVA: best-corrected visual acuity
CS: contrast sensitivity
AAO: American Academy of Ophthalmology

Ophthalmologists practicing for six or less years were more likely to have had a formal vision rehab rotation during their residency training compared to ophthalmologists practicing for seven or more years (*P* = 0.03). Ophthalmologists who completed a formal vision rehab rotation during residency reported greater confidence in their ability to appropriately identify patients who could benefit from vision rehab referral (*P* = 0.04) and referred patients for vision rehab at earlier visual acuity thresholds (*P* = 0.04) compared to ophthalmologists who did not complete a formal rotation during residency.

Ophthalmologists practicing for ten or less years were more likely to have had occupational therapists (OT) or certified low vision therapists (CLVT) as referral options at their residency institution compared to ophthalmologists practicing for over ten years (*P* = 0.01). However, access to OT and CLVT during residency did not significantly affect ophthalmologists’ vision rehab referral confidence or visual acuity thresholds.

Ophthalmologists practicing for over ten years were more likely to refer patients to vision rehab services in their own clinics (*P* = 0.001), but less likely to refer to other providers in their community (*P* = 0.03) compared to ophthalmologists practicing ten years or less. The clinical subspecialty of the ophthalmologists surveyed did not have a significant effect on vision rehab referral confidence or thresholds.

## Discussion

Current ACGME requirements allow for variable levels of vision rehab training among ophthalmology residency programs. Therefore, it is critical to analyze the effects of vision rehab training during residency on future referrals once trainees have graduated and entered clinical practice. This study specifically examines ophthalmologists’ confidence in identifying appropriate patients for vision rehab referrals and the visual criteria, including best-corrected visual acuity, used when referring. These metrics are important to characterize because early referral to vision rehab programs has been shown to improve outcomes.^8^

Approximately five years prior to this study, the Academy released a statement defining early referral for vision rehab services as the standard of care for patients with vision loss.^9^ Within this statement, the Academy made it clear that referral is indicated when a patient demonstrates best-corrected visual acuity of less than 20/40, scotomata, visual field loss, or contrast sensitivity loss which interferes with their daily activities.^9^ Early referral and treatment are vital to help prevent loss of reading ability, independence, mobility skills, and quality of life for those experiencing vision impairment.^9,10^ Our findings show that trainees who have graduated since the Academy’ s effort to promote early vision rehab referral are more likely to have had a vision rehab rotation during residency. These physicians are also making earlier referrals than their predecessors. This suggests that the Academy’ s efforts may be moving the needle in a positive direction for visually impaired patients. It also indicates that ophthalmology residents would benefit from more structured vision rehab training mandates.

The observation that internal vision rehab referrals are more likely to be made by ophthalmologists practicing for more than ten years may be due primarily to practice modality. Our survey did not ask about practice modality, but many vision rehab programs are found in academic settings. It is possible that longer-practicing ophthalmologists are more likely to be in settings which offer vision rehab services and therefore internal referrals are more common than to referring out for vision rehab services in this demographic.

There are several barriers for eligible patients to access vision rehab services; including lack of provider referrals, awareness of benefits, transportation, and financial resources.^11,12^ Expanding vision rehab training during residency can help alleviate some of this burden by preparing ophthalmologists to provide timely referrals and to educate patients about the availability and benefits of these services. Increased education and utilization will improve public awareness of vision rehab services, perhaps impacting downstream funding support and alleviating some of the cost-related concerns noted above.

Our findings lay the groundwork for a more comprehensive investigation of vision rehab referral patterns and training protocols among ophthalmologists. Our decision to recruit via email and social media solicitations may have caused the cohort to skew toward ophthalmologists earlier in their careers, as approximately 74% of subjects reported having been in practice for six years or less. A more formal survey of a larger and more representative cohort of practicing ophthalmologists, perhaps administered through the Academy itself, would be valuable. Another limitation of our study lies in the ability of physicians to accurately recall components of a training program which they may have completed over a decade ago. Pairing future studies with a formal survey of ophthalmology residency program directors may also help to corroborate responses and lead to more reliable conclusions. Finally, the temporal correlation between Academy efforts to promote early vision rehab referral and increasing residency exposure and referral confidence among ophthalmologists may not represent true causation.

## Data Availability

All data produced in the present study are available upon request.

## Acknowledgments

The study was supported in part by a Research to Prevent Blindness unrestricted grant to the Vanderbilt Eye Institute. The sponsor or funding organization had no role in the design or conduct of this research.

## Appendix

### Ophthalmology Low Vision Questionnaire

1. Did you have a formal low vision rehabilitation (VR) rotation during residency? Yes No
2. Were occupational therapists and/or certified low vision therapists available referral options at your residency institution? Yes No Unsure
3. What clinical findings would prompt you to refer to VR, if any? Please select all that apply.
  - Reduced best-corrected visual acuity
  - Visual field loss
  - Decreased contrast sensitivity
  - Other (describe):
4. At what best-corrected visual acuity level would you first consider referring a patient to VR? Please circle one, if applicable. 20/25 to 20/63 20/80 to 20/160 20/200 to 20/400 worse than 20/400
5. Since being in practice, have you referred patients to VR? Yes No
6. What VR resources do you present to visually impaired patients, if any? Select all that apply.
  - My clinical practice offers these services
  - State-run services
  - Non-governmental organizations
  - Community based providers
  - Online AAO resources
  - Other (describe):
7. How confident are you in your ability to appropriately identify patients who could benefit from VR, on a scale of 1 to 5? (1 = very confident, 5 = not at all confident) 1 2 3 4 5
8. Please indicate your subspecialty area of clinical practice within ophthalmology, if applicable:
9. How long ago did you complete your training? <3 years 3-6 years 7-10 years >10 years

